# Young people with obesity and rare disease – genotypes, phenotypes and healthcare use

**DOI:** 10.64898/2026.08.25.26361359

**Authors:** Chloe Chia, Kate Baker

## Abstract

Obesity is a significant public health concern. Early-onset obesity in the context of rare disease can reflect genetically-mediated pathology or elevated susceptibility through indirect mechanisms. Mapping the diverse characteristics and needs of young people with obesity in the rare disease population is a first step toward mechanistic and translational research. We carried out a retrospective comparative analysis of demographic, genotypic, phenotypic and health service utilisation data for young people with obesity (cases: n=500) and without obesity (controls: n=11,444) from the UK 100,000 Genomes Project rare disease cohort. Cases and controls were recruited prior to genomic diagnosis, across clinical disorder categories. We observed significant association between socioeconomic deprivation and obesity risk. Young people with obesity had significantly higher utilisations of acute care and mental health services, indicating an overall higher health burden. A curated panel of 519 candidate obesity-associated genes demonstrated aggregate association with obesity, although no single gene reached significance. Phenotypic comparison between cases and controls highlighted increased multi-organ and neurological system involvement, highlighting the overlap between neurodevelopmental and obesity risks. Within the case group, we conducted cluster analysis to identify early-onset obesity groups with different phenotypic profiles, potentially arising from different causal pathways - this identified six obesity subgroups of interest, with differing involvement of neurodevelopmental and other systems. Our study confirms that obesity co-occurs with a wide range of factors within the rare disease population, and is associated with significant physical and mental health needs, requiring holistic lifelong care.

## INTRODUCTION

Obesity, defined in children as a body mass index (BMI) at or above the 95^th^ centile, is a significant public health concern associated with negative health-related outcomes (1). Paediatric obesity usually arises from interactions between polygenic inheritance and environmental factors such as physical inactivity and diet (2–4). Less commonly, a specific underlying cause such as cerebral injury, endocrine dysfunction or rare genetic condition is diagnosed (1, 5). Obesity risk in the context of rare disease (RD) reflects a spectrum of association, ranging from high obesity penetrance via direct molecular mechanisms e.g. *LEP* and *MC4R* variants (6, 7), to elevated obesity risk via endocrine and neurobehavioural characteristics e.g. Prader-Willi syndrome (PWS) (8) and Bardet-Biedl syndrome (BBS) (9). Identifying a genetic cause of obesity can lead to initiation of targeted treatments: leptin replacement therapy can reduce body weight and improve endocrine function (10); growth hormone treatment in PWS leads to significant improvements in BMI and cognition (11).

Current genomic diagnostic yields for individuals with suspected syndromic obesity approach 20% (7, 12, 13). However, the proportion of individuals with RD who develop early-onset obesity has not been assessed. Moreover demographic, genetic and phenotypic predictors of obesity in the context of RD have not been comprehensively investigated. Pinpointing individuals at risk of obesity at the time of RD diagnosis could facilitate proactive management to improve long-term health outcomes.

Toward these goals, this study investigated early-onset obesity within the 100,000 Genomes Project (100KGP) (14), which applied whole genome sequencing to individuals with suspected RD (15, 16). Our first objective was to compare case (early-onset obesity present) and control (early-onset obesity absent) groups with respect to demographics, rare variant burdens, phenotype catalogues and healthcare use (as a proxy for multi-morbidity). Our second objective was to perform phenotypic clustering within the case group to delineate rare obesity sub-populations, informing future mechanistic research and stratified care.

## MATERIALS AND METHODS

### Cohort Curation

This project was approved by the 100KGP research registry. Analyses were carried out within the GEL Research Environment, and queries performed using the LabKey Python API. General inclusion criteria were (i) proband status, (ii) active consent status, (iii) phenotypic-karyotypic sex concordance, and (iv) age <18 years at point of recruitment to 100KGP (conservatively estimated using a year of birth cut-off of 2003, given last study recruitment date 29 Dec 2020).

Cases were defined by the presence of an obesity-related term in at least one of three sources: (A) clinician-reported Human Phenotype Ontology (HPO) terms at recruitment (17); (B) ICD-10 codes in linked NHS Hospital Episode Statistics (HES) (18); (C) recruitment to 100KGP via the normalised disease subgroup “obesity syndromes (10973)”. See Table S1 for HPO and ICD-10 code lists applied for case definition. Controls were all rare disease cohort participants fulfilling the general inclusion criteria but not meeting any obesity-related criterion.

### Demographics

Current age was calculated from year of birth at the time of data extraction (rather than age at recruitment, to account for obesity-related ICD-10 codes and healthcare utilisation after 100KGP enrolment). Ethnicity was derived hierarchically using genomic ancestry information and, if unavailable, self-reported ethnicity. Socioeconomic status was approximated via HES Index of Multiple Deprivation (IMD) score.

### Genetic Characterisation

Rare genomic variants had been identified via the automated GEL pipeline and categorised via the Tiering system (19). We compared the proportion of cases and controls with Tiered variant(s) considered potentially diagnostic, and compared total rare variant burdens between groups. To assess whether the catalogue of specific rare variants differs between cases and controls, group-level gene burden panels were compiled (number of unique participants with Tier 1 or 2 variants in each gene) and compared via panel-level Fisher’s exact test (two-tailed 519 x 1 x 1). Next, we performed gene-level enrichment analysis across all 1916 genes identified in both groups, using two-tailed Fisher’s exact tests with Benjamini-Hochberg (BH) correction.

### Phenotypic Characterisation

Each participant’s complete list of HPO annotations was retrieved. To focus on co-occurring phenotypes, obesity-related HPO terms (Table S1) were excluded. Raw HPO annotations were parsed and propagated to include all parent terms up to the root term “HP:0000118 - Phenotypic abnormality” using PyHPO 4.0.0 (17). Differences in median number of top-level and raw terms per participant were assessed using Mann-Whitney U tests. We then compared (i) top-level terms (classified by body system), and (ii) the full propagated set including raw and intermediate terms, using two-proportions z-tests with BH correction.

### Health service use

Emergency care utilisation was assessed using HES A&E records and the Emergency Care Data Set (ECDS), reflecting NHS transition from April 2020. A&E visits were identified within a five-year observation window (31/07/2017-31/07/2022), defined from the latest available encounter date in the cohort, with visits de-duplicated by participant ID and visit date. Utilisation rates were modelled as visit counts per person-year. Given non-normal distributions confirmed by Kolmogorov-Smirnov testing, differences in median visit counts were assessed using Mann-Whitney U tests, and incidence rate ratios (IRRs) were calculated. Where available in ECDS records, SNOMED-CT chief complaint and primary diagnosis codes were extracted, with visits lacking clinical codes excluded from presentation-level analyses. The 10 most frequent A&E presentation reasons were compared between groups using chi-square or Fisher’s exact tests with BH correction, treating each visit as a separate event to quantify overall emergency care burden.

Admitted Patient Care (APC) utilisation was analysed using HES APC records following the same framework as emergency care analyses, including calculation of admission rates per person-year and identification of the 10 most common primary diagnoses. In addition to per-admission analyses capturing overall service burden, a secondary patient-level analysis was performed, counting unique primary diagnosis per participant across the five-year window.

NHS mental health services use was identified through the Mental Health Services Data Set (MHSDS) master patient index. Referral reasons and diagnoses were extracted from linked service referral, diagnosis, and curated assessment records, with ICD-10 mental and behavioural disorder codes (F00-F99) propagated to higher-level categories using the simple_icd_10 Python library v2.1.1 (20). Mental health contact counts and rates per person-year were derived from curated community and care contact records with deduplication. Shapiro-Wilk testing confirmed non-normality - between-group differences were assessed using Mann-Whitney U tests and IRR analyses.

### Phenotypic clustering

To identify phenotypically distinct subgroups within the case cohort, we applied a semantic similarity-based clustering approach (21). To improve the signal-to-noise ratio, patients with fewer than 6 (based on Q1=5 + 1) raw terms were removed. Raw HPO annotations of the remaining cases were parsed and propagated. To mitigate bias due to variability in term count, HPO term list lengths were equalised by capping each case’s term list at the upper outlier threshold (Q3□+□1.5□×□IQR). To focus on high information content terms, modifier and obsolete terms were removed, and parent terms excluded when a more specific descendant was present. Pairwise semantic similarity between participants was computed using the *funSimAvg* method (21). Similarity scores were converted to distances and clustered hierarchically using Ward’s method (22). Cluster number was chosen by manual dendrogram inspection and 0.95 threshold by relative dendrogram height on the normalised linkage distance scale. Clusters with fewer than five cases were dropped.

To map the phenotypic features driving cluster membership, the proportion of participants with each propagated HPO term in each cluster was compared against all other clusters’ using Fisher’s exact tests with BH correction. To explore the genetic architecture of each phenotype cluster, we compiled genes harbouring Tier 1 or 2 variants in at least one participant within each cluster. Gene-level overlap between clusters was quantified using pairwise Jaccard indices (size of the intersection divided by size of the union of gene sets for each cluster pair). Cluster pairs with no shared genes were assigned a Jaccard index of 0. Functional ontology enrichment of gene sets observed within each cluster were visualised using ShinyGO (https://bioinformatics.sdstate.edu/go/).

## RESULTS

### Cohort ascertainment

500 cases and 11,444 controls were identified, corresponding to 4.2% within-cohort recorded prevalence for early-onset obesity. Figure 1A illustrates case ascertainment, noting ICD-10 codes as most frequent source, and limited overlap between sources. Table S2 lists clinician-reported HPO ascertainment terms. Figure 1B shows the distribution of 100KGP normalised disease recruitment categories across case and control groups. “Neurological and neurodevelopmental disorders” was the most frequent category in both groups. A higher proportion of cases than controls were recruited for endocrine disorders and multi-system disorders, whilst a higher proportion of controls were recruited to single-system categories. Early-onset obesity was present in 4.4% of rare disease cohort members recruited for neurological disorders and 17.8% recruited for endocrine disorders (Figure S1).

**Figure 1:**
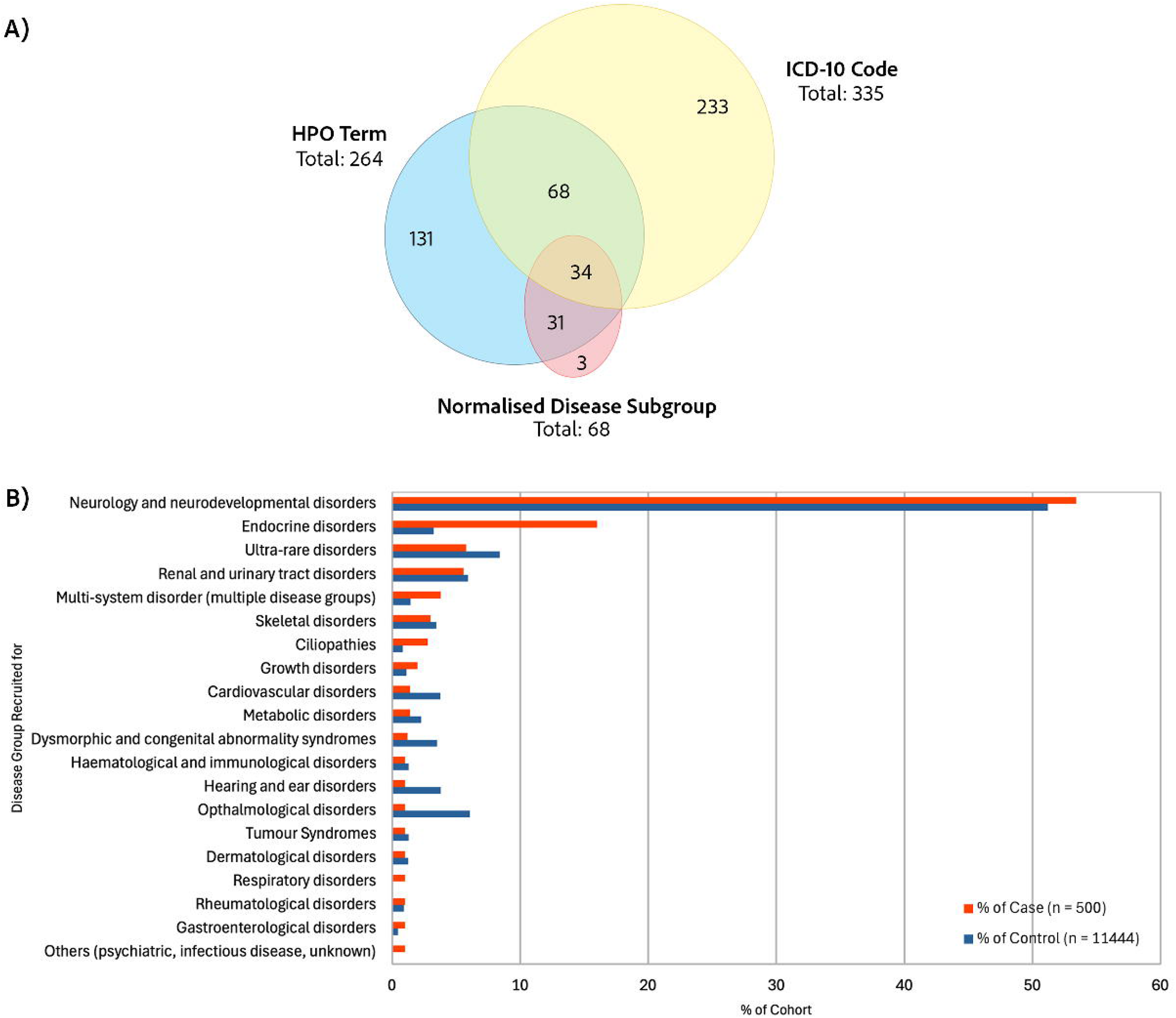
Case ascertainment sources and 100KGP disease recruitment categories. A) Venn diagram showing the number of cases identified to have/had obesity via i) clinician-reported HPO term at 100KGP study recruitment, ii) ICD-10 code in NHS health records, iii) 100KGP normalised disease recruitment category “obesity”. Some cases had obesity-related codes in multiple sources. B) Distribution of cases and controls across disease recruitment categories. Participants recruited into multiple categories are grouped into “multi-system disorders”. Categories with fewer than 5 individuals have been masked (rounded up to 5) to protect privacy.

### Demographics

Table 1 and Figure S2 describe groups’ demographics. Cases’ age distribution was significantly shifted toward being older (at time of data analysis). No significant difference in sex or ethnicity composition was found. Cases exhibited a significantly lower median IMD quintile, reflecting a greater representation of cases within more deprived quintiles.

**Table 1:** Demographics of case and control groups.

|  | Case | Control | Comparison |
| --- | --- | --- | --- |
| **Total number of participants** | 500 | 11 444 |  |
| **Age** |  |  |  |
| **Median / years (IQR)** | 16 (13 – 19) | 14 (11 – 17) | $p = 4.57e-21$ |
| **Current age band** |  |  |  |
| <10 years N (%) | 16 (3.2%) | 1301 (11.3%) |  |
| 10 – 14 years N (%) | 160 (32%) | 5000 (43.7%) |  |
| 15 – 19 years N (%) | 220 (44%) | 3602 (31.5%) |  |
| >20 years N (%) | 104 (20.8%) | 1541 (13.5%) |  |
| **Sex** | | | $\chi^2 p = 0.925$ |
| Male N (%) | 289 (57.8%) | 6651 (58.1%) |  |
| Female N (%) | 211 (42.2%) | 4793 (41.9%) |  |
| **Ethnicity** | | | $\chi^2(4) p = 0.551$ |
| White N (%) | 372 (74.4%) | 8857 (77.4%) |  |
| Asian N (%) | 76 (15.2%) | 1533 (13.4%) |  |
| Black N (%) | 14 (2.8%) | 299 (2.6%) |  |
| Mixed N (%) | 22 (4.4%) | 394 (3.4%) |  |
| Unknown/Other N (%) | 16 (3.2%) | 361 (3.2%) |  |
| **Index of Multiple Deprivation** |  |  |  |
| **IMD Median / quintile (IQR)** | | | $p = 1.33e-04$ |
| 1 - Most deprived N (%) | 143 (28.6%) | 2518 (22%) |  |
| 2 N (%) | 106 (21.2%) | 1990 (17.4%) |  |
| 3 N (%) | 75 (15%) | 1743 (15.2%) |  |
| 4 N (%) | 55 (11%) | 1585 (13.8%) |  |
| 5 - Least deprived N (%) | 62 (12.4%) | 1700 (14.9%) |  |
| Unknown N (%) | 59 (11.8%) | 1908 (16.7%) |  |

### Genomic characteristics

4256 Tier 1 or 2 variants were identified in 519 genes within 349 case participants. 89,801 Tier 1 or 2 variants were identified across 1889 genes in 7303 controls. Cases were more likely than controls to carry at least one Tier 1 or 2 variant (69.8% vs 63.8%), representing a modest increase in rare variant burden (Fisher’s exact test, *p = 0.0066;* OR 1.31, 95% CI 1.08–1.59). Diagnostic yield, defined by 100KGP as family case solved, was similar between cases (19.8%, n=99) and controls (20.9%, n=2386; Fisher’s exact test, *p = 0.613)*.

To evaluate whether rare, tiered variants in genes identified in cases were collectively enriched versus controls, we performed a panel-level analysis. Testing of this 519 gene set (df = 1) confirmed collective enrichment (OR = 2.26, *p = 1.86e-17)*, indicating that cases were more than twice as likely as controls to harbour a Tiered variant within this panel. Gene-level tests were performed on all case and control genes (n=1916 after deduplication). No statistically significantly enriched or depleted genes were identified. Table S3 lists genes with p_raw_ <0.01. The five genes with the strongest signal *(p<1.0)* enriched amongst cases were *FLNB, SCN4A, SIL1, MC4R,* and *BBS10*.

### Phenotypic associations

HPO terms per patient (Figure S3A) was skewed towards a higher number of raw terms for cases (median = 7, IQR 5-11) than controls (median = 7, IQR 4-10) *(p = 4.27e-03).* Using top-level terms to represent affected organ systems, there was a significant increase in number of systems affected in cases (median = 4, IQR 2-6) versus controls (median = 3, IQR 2-5) *(p = 1.48e-06)* (Figure S3B). Together, these results suggest increased phenotypic complexity amongst cases.

The percentage occurrence of top-level terms in each group was compiled (Table S4). Figure 2A shows the proportion of patients with abnormalities in each organ system in each group. While the majority of patients in both groups had nervous system abnormalities, this was further enriched in patients with obesity (Δ *= +16.2%, p_adj_ = 2.77e-13).* Other significantly enriched top-level terms include endocrine, growth, genitourinary and limb abnormalities. “Abnormality of the cardiovascular system” was the only significantly reduced term within the obesity case group (Δ *= -4.92%, p_adj_ = 0.0163)*.

**Figure 2:**
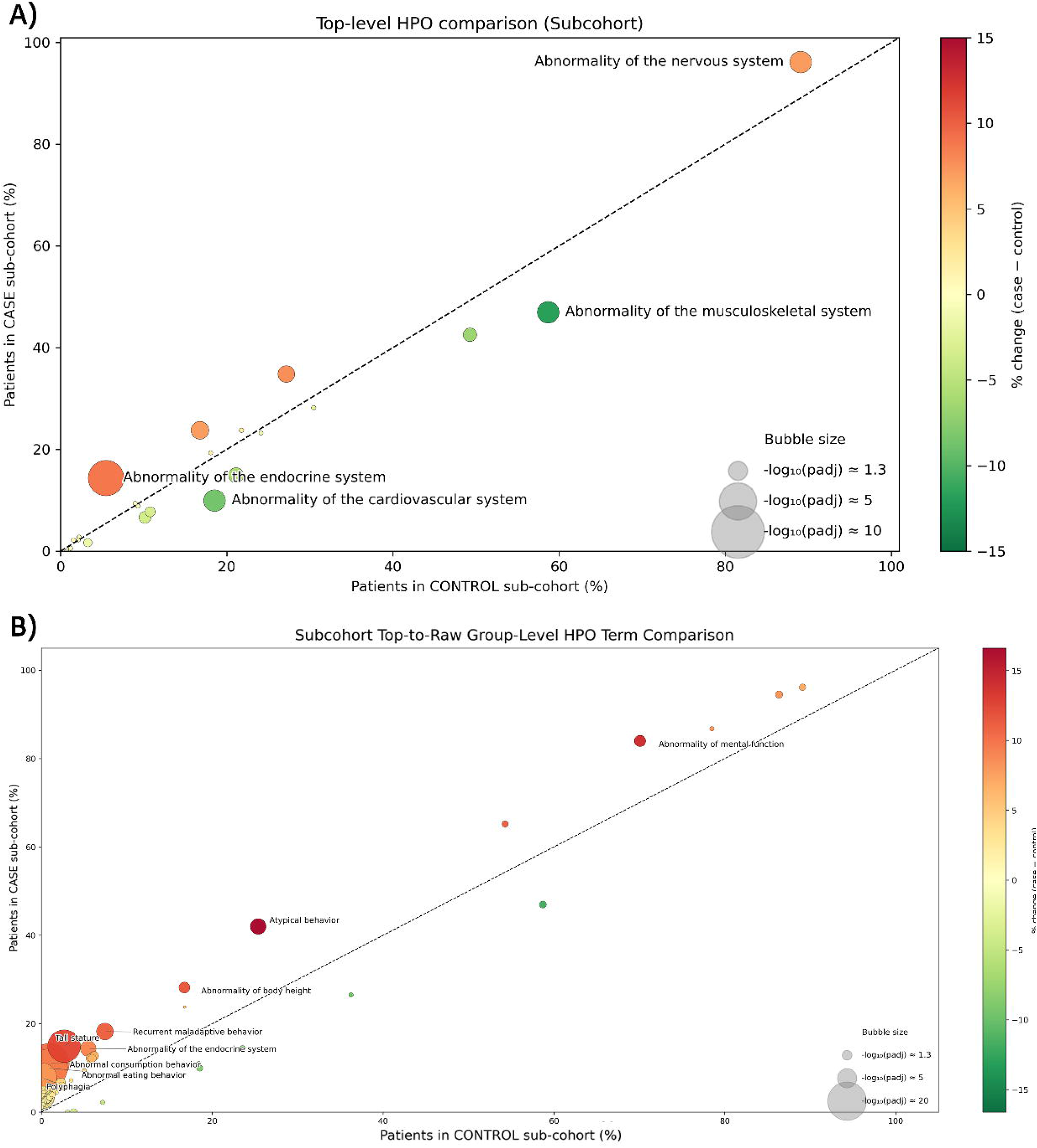
Frequency of HPO terms within case and control groups. A) Top-level HPO term comparison between case and control groups. Dots represent the proportion of cases (y-axis) and controls (x-axis) with significantly enriched *(|*Δ*|>3%, p_adj_ <□0.05)* top-level HPO terms, scaled for between-group difference. B) Full propagated HPO term comparison between case and control groups. Only terms present in at least 15% of either group are labelled.

We additionally compared the full set of child terms descended from top-level terms (Figure 2B; Table S5). Most terms clustered in the bottom left of the plot, indicating low frequencies in both groups. This includes phenotypes significantly enriched amongst cases, such as “polyphagia” (5.8% cases, 0.05% controls) and “sleep apnoea” (3.8% cases, 0.72% of controls). Additionally, phenotypes relating to cognitive function were significantly enriched in cases, including “intellectual disability” (Δ *= +14.9%, p_adj_ = 8.42e-09)* and “atypical behaviour” (Δ *= +15.8%, p_adj_ = 7.57e-18)*.

Seven significantly enriched or reduced terms did not belong to any previously highlighted top-level system, including: “hypertonia”, “abnormal appendicular skeleton morphology”, and “macrocephaly”. These are all “abnormalities of the musculoskeletal system”. The lack of detection of top-level significance was likely due to opposing directions of enrichment – while skeletal abnormalities were enriched in obesity patients, muscular abnormalities were more common amongst controls. In this analysis, cardiovascular abnormalities were not significantly reduced after multiplicity correction.

### Healthcare use

1800 and 34,427 A&E visits were recorded for 500 cases and 11,444 controls. Figure S4A shows visits per participant within both groups. Most patients had fewer than five A&E visits in the five-year period. Cases had a significant increase in A&E attendance (median = 2, IQR 0 – 4) compared to controls (median = 1, IQR 0 – 4) *(p = 5.84e-04)*. Mean A&E visits was 0.72/person-year in cases and 0.60/person-year in controls, giving an IRR of 0.84 (95% CI 0.80 – 0.88, *p = 1.12e-13)*.

Of 1460 case and 28,217 control A&E visits queried, 1229 (84.2%) case and 24,509 (86.9%) control visits had chief complaint information available; 1137 (77.9%) case and 21,915 (77.7%) control visits contained primary diagnosis information. Figure S4 shows each group’s top 10 chief complaints and primary diagnoses. The odds ratio of each condition and Chi-square test results with BH correction are available in Tables S6 and S7. The most common chief complaints were similar, with 9 of 10 categories overlapping across groups. However, cases had significantly higher odds of presenting with “injury of lower limb” (OR: 1.82, *p_adj_ = 1.25E-10)* and “abdominal pain” (OR: 1.37, *p_adj_ = 0.023)*, and lower odds of presenting with “seizure”, “fever”, “injury of head”, and “dyspnoea”. The spectrum of primary diagnoses in both groups was also similar, with an overlap of 8 of 10 categories and the same top 3 most frequent diagnoses. Obesity patients were significantly more likely to present with “sprain of ankle” (OR: 2.72, *p_adj_ = 1.50E-10)* and “superficial injury of foot” (OR: 1.93, *p_adj_ = 0.0227)*.

3167 case and 47,181 control inpatient admissions were observed within the five-year window (Figure S5A). Most participants had fewer than 10 admissions, with a small subset having more than 40. Cases had a significantly higher number of admissions (median = 2, IQR 0 - 5) than controls (median = 1, IQR 0 - 4) *(p = 1.56e-06).* The mean person-year-adjusted rate of admitted patient care in cases and controls was 1.27/py and 0.82/py respectively, giving a control-case IRR of 0.65 (95% CI 0.80 - 0.88, *p <1.00e-13)*.

ICD-10 codes were available for all admissions in this dataset. We compared the top 10 most frequent primary diagnoses in both groups (Figure S5B), finding an overlap of only five categories. Cases were significantly more likely to be admitted for chronic kidney disease (CKD) stage 5 (OR: 5.79, *p_adj_ = <0.001)*, aplastic anaemia (OR: 5.90, *p_adj_ = 4.92E-60)*, sleep apnoea (OR: 1.55, *p_adj_ = 2.43E-03)*, precocious puberty (OR: 6.11, *p_adj_ = 4.33E-35)*, and hypopituitarism (OR: 3.11, *p_adj_ = 1.28E-11)*. Conversely, they had significantly lower odds of being admitted for respiratory infections, viral infections, unspecified epilepsy, and holiday relief care. Unique diagnosis codes per participant (Figure S5C) indicated that neither CKD stage 5 or aplastic anaemia were top ten admission reasons for either group, indicating that cohort-level results were likely driven by small case numbers with repeated admissions. Cases were more frequently admitted for sleep apnoea (OR: 2.16, *p_adj_ = 1.42E-04)*, hypopituitarism (OR: 3.60, *p_adj_ = 5.29E-06)* and, notably, obesity itself (with lack of additional specification).

149 (29.8%) cases and 2290 (20%) controls were known to NHS Mental Health Services (MHS). The number of care encounters for each participant (31/07/2017-31/07/2022) was compiled as a proxy measure of MHS utilisation (Figure 3A). Most patients had fewer than five care encounters, with a subset having 30 or above. Cases had a significant increase in MHS encounters (median = 2, IQR 0 - 10) compared to controls (median = 1, IQR 0 - 6) *(p = 0.0093)*. MHS referral reasons were retrieved for 76 case and 862 control patients (Figure 3B) - the top five referral reasons overlapped completely but differed in order between groups. Code “17 - neurodevelopmental conditions” (retired 1 April 2018) combined with the replacement code “24 - neurodevelopmental conditions, excluding autism” (23) were the commonest referral reason across both groups (36% and 34%). Conduct disorders accounted for 25% of referrals for cases and 13.8% for controls.

**Figure 3:**
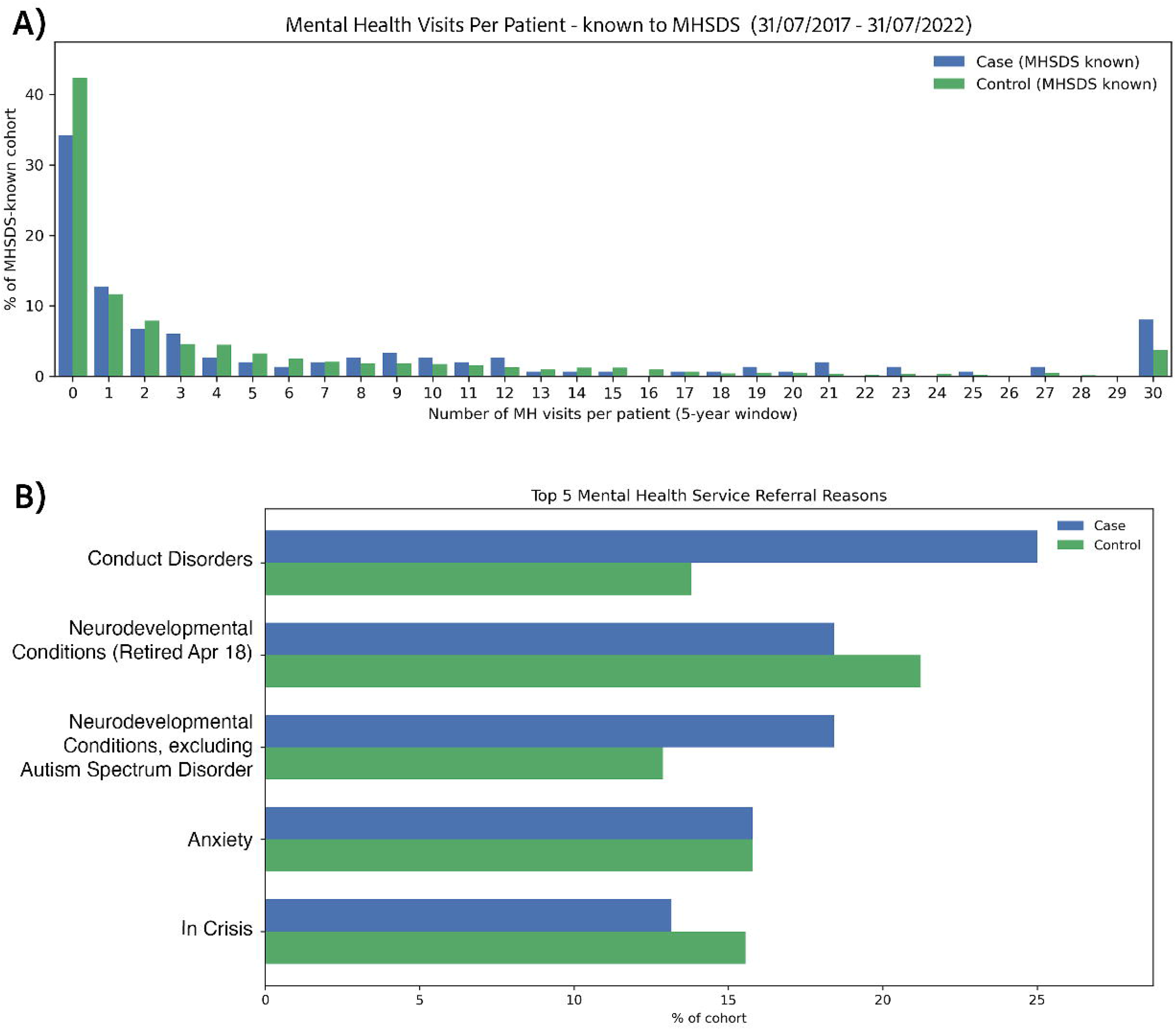
Mental health service use in cases and controls. Distribution of mental health encounter visit numbers and service referral reasons of participants known to NHS Mental Health Services (31/07/2017 – 31/07/2022). A) Percentage distribution of mental health encounters per participant. Participants with more than 30 mental health visits are grouped in the 30 bin. B) Top 5 NHS mental health service referral reasons amongst participants with known referral reasons to NHS Mental Health Services.

### Phenotype clustering of early-onset obesity cases

Given the extent of genotypic and phenotypic heterogeneity within obesity-associated rare disease, we investigated whether data-driven sub-groups can be identified based on phenotypic similarity. Within the obesity case group, 15 phenotypic similarity clusters were identified from manual dendrogram inspection (Figure 4A), and 11 clusters retained with greater than five patients. Diagnostic plots show high intra and inter-cluster variability (Supplementary Figure 6) indicating HPO list length is not the key factor driving cluster formation. Inter-cluster analysis revealed 20 HPO terms significantly enriched in any one cluster compared to the remainder, where OR>1 and p_adj_ <0.05 (Figure 4B; Table S7). These terms are represented across six clusters: 1, 2, 6, 7, 9, and 10.

**Figure 4:**
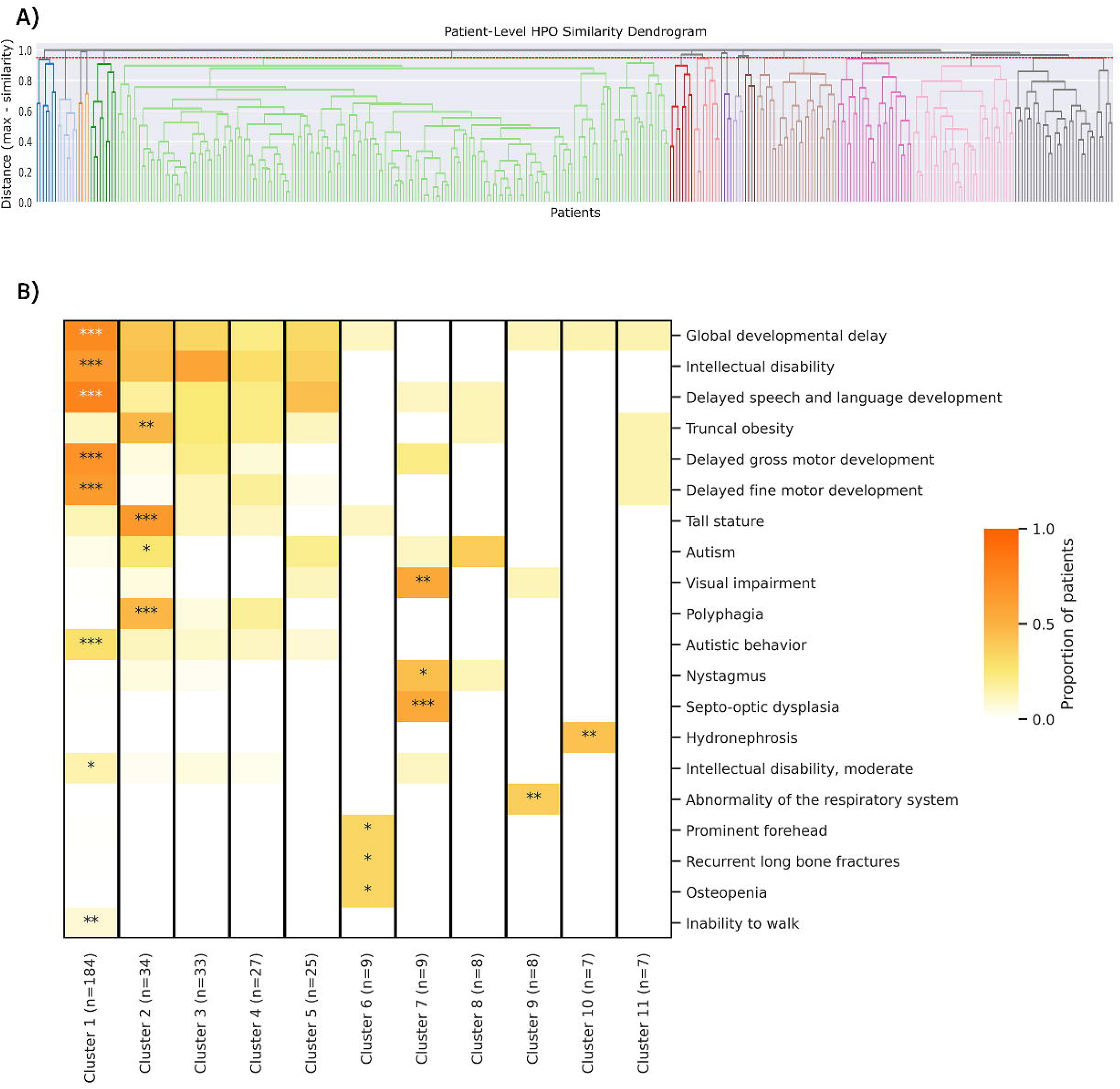
Obesity case group - phenotypic similarity clusters. A) Case-Level Dendrogram B) Cluster-level proportion occurrence of significantly enriched (OR > 1 and p_adj_ < 0.05) HPO terms (Fisher’s exact test with Benjamini-Hochberg (BH) correction). *** = p_adj_ <0.001, ** = p_adj_ <0.01, * = p_adj_ <0.05.

Cluster 1, by far the largest (n=184), is defined by neurodevelopmental abnormalities such as global developmental delay and ID. Although cluster 2 is also characterised by neurodevelopmental difficulties, additional hallmarks include truncal obesity, tall stature, polyphagia and autism; motor and communication delays were less frequently reported. Clusters 6 to 11 each contained a small number of participants, sharing phenotypes that were rarer within the cohort overall. Clusters 6, 7, 9, and 10 are characterised by skeletal, optic, respiratory, and renal abnormalities respectively. The list of genes harbouring Tier 1 or 2 variants in at least one participant per cluster was compiled. The number of genes per cluster ranged from 6 to 269. Both intra- and inter-cluster genetic heterogeneity were observed – gene sets identified within each phenotype cluster showed little overlap across clusters (all pairwise Jaccard index values <0.1) (Table S8). Visualisation of gene ontology enrichment networks within clusters (Figure S7) indicated that Clusters 1 and 2 are functionally diverse, encompassing neuronal excitability genes and chromatin-associated genes, as expected for rare neurodevelopmental conditions. Cluster 7, a small N cluster defined phenotypically by visual impairments, demonstrates strong representation of a chromatin-associated network only. Remaining clusters did not demonstrate gene ontology enrichments.

## DISCUSSION

In this study, we investigated demographic, genomic, and phenotypic risk factors associated with early-onset obesity in the RD cohort, and documented multi-morbidity via health service use. We explored phenotypic heterogeneity within RD-obesity, with a view to future mechanistic and therapeutic studies.

Greater deprivation amongst obesity cases reflects the wider UK population – at age six, deprived children are 17% more likely to be obese than their peers (24), mediated by restricted access to healthy food and physical activity (25, 26). Our results indicate these factors are also important for young people with RD, highlighting the need for access to population-level interventions for those with complex needs. Older age of the RD-obesity case group suggests that deprivation contributes to the emergence of obesity over time (27, 28).

There was no substantial impact of early-onset obesity on likelihood of RD genomic diagnosis (noting that diagnostic yield relates to the 100KGP recruitment indication, not necessarily obesity). However, we observed a modestly increased rare variant burden in cases, and collective enrichment in panel-level rare variant analysis. This suggests that the excess rare variant burden in cases is concentrated within a subset of genes. This enrichment was not driven by statistically significant effects for individual genes, likely due to limited power and stringent multiplicity correction, rather than an absence of true genetic effects. Meta-analyses combining large cohorts such as UK Biobank (29) may overcome this, however translating group-level enrichment to individual prediction of obesity risk is likely to remain challenging.

Several candidate rare genomic risk factors for obesity were observed. *SCN4A* and *FLNB* have the highest case-control ORs of 69 and 6. However, these variants were diagnostic for only one of three patients with *SCN4A* variants, and no patients with *FLNB* variants. Limited literature is available regarding their associations with obesity, beyond a *SCN4A* mouse model study (30) and a study of monozygotic twins with discordant BMI that investigated *FLNB* expression in skeletal muscle proteomes (31). Interestingly, both studies linked variants in these genes to reduced body weight. *BBS10* and *MC4R* have established links to obesity (32, 33). Other observed candidate genes include *CHAMP1, NEXMIF* and *TGFB2.* In a case series of patients with *CHAMP1*-associated disorder, 8% presented with obesity, although others were underweight (34, 35). *NEXMIF* is associated with an X-linked neurodevelopmental disorder (36), with obesity reported as an additional feature (37, 38). While *TGFB2* has not previously been associated with obesity, it belongs to the TGF-β pathway which exerts regulatory effects on adipogenesis (39).

Considering phenotypic risk factors for obesity, we first noted that obese cases have higher phenotypic burdens, reflecting increased RD complexity beyond obesity. Previous reviews of syndromic obesity have highlighted its high phenotypic variability (1, 7, 40). Consistent with this, our term frequency analyses revealed multi-organ involvement within the case group. This increased clinical complexity was also reflected in healthcare utilisation patterns, corroborating findings from the general paediatric population. A large meta-analysis reported higher healthcare utilisation among obese children, including a 34% relative increase in A&E visits (35). The association between obesity and lower extremity injuries observed in our study is well established; a large U.S. study demonstrated increased risks of fractures, sprains, and musculoskeletal pain in children with obesity, independent of demographic factors (41). This underscores the importance of injury prevention strategies alongside promotion of physical activity in the RD population.

The most obvious phenotypic difference between case and control groups was the higher prevalence of complex neurodevelopmental difficulties, encompassing both intellectual disabilities and social-emotional-behavioural problems. Further research is needed to untangle in what circumstances neurodevelopmental phenotypes are causes, correlates or consequences of obesity. Phenotype descriptors such as “recurrent maladaptive behaviour” could encompass inflexible or compulsive behaviours that lead to weight gain, or represent responses to obesity and food-related stress (e.g. insatiable hunger, frustration, low self-esteem). Analysis of mental health service use data indicated that, for a proportion of cases within the analysed cohort, these difficulties are sustained and disruptive, escalating to conduct disorders. Indeed, in our study, mental health referrals were observed in 29.8% of young people with obesity compared with 20% of controls. This difference was unexpected given the RD background of the control cohort, and suggests an additional mental health burden associated with obesity. Longitudinal time-course HPO analyses that track the developmental sequence of phenotype emergence (accounting for heterogeneous rare aetiology) may provide insights with therapeutic relevance.

We carried out phenotypic cluster analysis as a step toward better understanding the heterogeneity of RD presentations associated with obesity. This revealed large clusters characterised by neurodevelopmental features, alongside smaller clusters representing more distinct phenotypic profiles. Notably, even within the dominant neurodevelopmental clusters, differences in cognitive and growth-related features indicate that obesity in RD is not phenotypically uniform but comprises distinguishable neurobehavioural sub-profiles, worthy of in-depth investigation in future studies. Phenotypic co-occurrence patterns within small clusters may reflect more direct mechanistic pathways (e.g. septo-optic dysplasia associated with hypothalamic dysfunction), clinical risk interactions with obesity (e.g. recurrent fractures risks, respiratory complications). This may have implications for personalised management, by proactively implementing preventive or monitoring measures. Analysis of genetic variants present within each cluster revealed substantial genetic heterogeneity, but limited overlap between clusters. This suggests that genetic factors and developmental mechanisms may contribute to phenotype differences between clusters.

Overall, our study highlights the complexity of rare developmental conditions associated with obesity, and catalogues genetic and psychosocial factors that may contribute to obesity risk. It also reiterates the need to consider the association between obesity and RD as a spectrum, rather than a dichotomy of syndromic or non-syndromic. Meeting the multi-disciplinary support needs for young people with RD-obesity requires enhanced integration of physical health, mental health and family support services – whilst such integration is costly, it may have long-term benefits that should not be ignored. Residual confounding due to differences in age and socioeconomic deprivation between groups cannot be excluded in our comparisons of healthcare utilisation. Nevertheless, the present findings reflect the real-world physical and mental healthcare burden experienced by children with RD-obesity – a socioeconomically disadvantaged population for whom cumulative health service utilisation is clinically and economically meaningful. This is especially so because obesity is a recognised risk factor for cardiometabolic disease, and increasingly for adverse brain health outcomes - early accumulation of multimorbidity can increase lifetime risk of dementia (42). Early-onset obesity may amplify existing vulnerabilities in individuals with RD. In populations already exposed to social and medical health inequalities, this may contribute to a cascade of risk across the lifespan, reinforcing the importance of early intervention.

## Data Availability

All data produced in the present work are contained in the manuscript

https://www.genomicsengland.co.uk

## DATA AVAILABILITY

Data from the National Genomic Research Library (NGRL) used in this research are available within the secure Genomics England Research Environment. Access to NGRL data is restricted to adhere to consent requirements and protect participant privacy. Data used in this research include rare disease participant phenotype, diagnostic status, and tiered variant datasets; linked NHS Hospital Episode Statistics (HES) A&E records, NHS Emergency Care Data Set (ECDS), and NHS Mental Health Services Data Set (MHSDS). Access to NGRL data is provided to approved researchers who are members of the Genomics England Research Network, subject to institutional access agreements and research project approval under participant-led governance. For more information on data access, visit: https://www.genomicsengland.co.uk/research.

## CODE AVAILABILITY

Code used for HPO phenotype analysis and figure generation (methods section 2.4) were adapted from previous work by Alice Smail (43), and can be found here: https://github.com/alicesmail12/HPOAnalysis.

## ACKNOWLEDGEMENTS

We gratefully acknowledge the participants of the National Genomic Research Library (NGRL), whose contributions made this research possible. Secure access to the NGRL under project ID 1251 was provided by Genomics England, which delivers the NGRL in partnership with NHS England, and is wholly owned by the UK Department of Health and Social Care. The NGRL contains participants’ health data collected by the NHS as part of their care, along with samples and data from their participation in research, for which fully informed consent has been obtained. This includes genomic and clinical data provided through the NHS Genomic Medicine Service, as well as data obtained through research studies, including the 100,000 Genomes Project and the Generation Study, both of which are delivered in partnership with the NHS, and from other research cohorts involving external collaborators. Analysis concepts, design, interpretation and manuscript preparation were funded by the Medical Research Council (MC_UU_00030/3).

## AUTHOR CONTRIBUTION STATEMENT

Both authors conceived and designed the study. CC carried out all data analyses and wrote the manuscript. Both authors revised the manuscript.

## ETHICAL APPROVAL

This study involves data previously collected for human participants, which was approved for Genomics England: HRA Committee East of England–Cambridge South (REC ref: 14/EE/1112). Participants gave informed consent to participate in the study before taking part.

## REFERENCES

1. Carvalho LML, Jorge LDAA, Bertola RD, Krepischi VCA, Rosenberg C. A Comprehensive Review of Syndromic Forms of Obesity: Genetic Etiology, Clinical Features and Molecular Diagnosis. Current Obesity Reports. 2024;13(2):313–37.

2. Lecroy NM, Kim SR, Stevens J, Hanna BD, Isasi RC. Identifying Key Determinants of Childhood Obesity: A Narrative Review of Machine Learning Studies. Childhood Obesity. 2021;17(3):153–9.

3. Nau C, Ellis H, Huang H, Schwartz SB, Hirsch A, Bailey-Davis L, et al. Exploring the forest instead of the trees: An innovative method for defining obesogenic and obesoprotective environments. Health & Place. 2015;35:136–46.

4. Gray CJ, Schvey AN, Tanofsky-Kraff M. Demographic, psychological, behavioral, and cognitive correlates of BMI in youth: Findings from the Adolescent Brain Cognitive Development (ABCD) study. Psychological Medicine. 2020;50(9):1539–47.

5. Farooqi SI, O’Rahilly S. Genetics of Obesity in Humans. Endocrine Reviews. 2006;27(7):710–8.

6. Farooqi SI, O’Rahilly S. Mutations in ligands and receptors of the leptin–melanocortin pathway that lead to obesity. Nature Clinical Practice Endocrinology & Metabolism. 2008;4(10):569–77.

7. Kaur Y, Souza DJR, Gibson TW, Meyre D. A systematic review of genetic syndromes with obesity. Obesity Reviews. 2017;18(6):603–34.

8. Muscogiuri G, Barrea L, Faggiano F, Maiorino IM, Parrillo M, Pugliese G, et al. Obesity in Prader–Willi syndrome: physiopathological mechanisms, nutritional and pharmacological approaches. Journal of Endocrinological Investigation. 2021;44(10):2057–70.

9. Beales PL, Cetiner M, Haqq AM, Miller J, Shoemaker AH, Valverde D, et al. Hyperphagia in Bardet-Biedl syndrome: Pathophysiology, burden, and management. Obes Rev. 2025;26(7):e13915.

10. Paz-Filho G, Mastronardi CA, Licinio J. Leptin treatment: Facts and expectations. Metabolism. 2015;64(1):146–56.

11. Passone BGDC, Franco RR, Ito SS, Trindade E, Polak M, Damiani D, et al. Growth hormone treatment in Prader-Willi syndrome patients: systematic review and meta-analysis. BMJ Paediatrics Open. 2020;4(1):e000630.

12. Partenope C, Monteleone G, Andorno S, Petri A, Prodam F, Bellone S, et al. Towards a genetic obesity risk score in a single-center study of children and adolescents with obesity. Scientific Reports. 2025;15(1).

13. Dubern B, Mosbah H, Pigeyre M, Clément K, Poitou C. Rare genetic causes of obesity: Diagnosis and management in clinical care. Annales d’Endocrinologie. 2022;83(1):63–72.

14. The National Genomic Research Library, Genomics England (2024). 10.6084/m9.figshare.4530893.

15. The 100 GPPI. 100,000 Genomes Pilot on Rare-Disease Diagnosis in Health Care — Preliminary Report. New England Journal of Medicine. 2021;385(20):1868–80.

16. Mark C, Jim D, Martin D, Leila E, Tom F, Sue H, et al. National Genomic Research Library v5.1, Genomics England. doi:10.6084/m9.figshare.4530893/7. 2020.

17. Gargano MA, Matentzoglu N, Coleman B, Addo-Lartey EB, Anagnostopoulos AV, Anderton J, et al. The Human Phenotype Ontology in 2024: phenotypes around the world. Nucleic Acids Res. 2024;52(D1):D1333–d46.

18. Organization WH. International statistical classification of diseases and related health problems (11th edition). 2019.

19. England G. Rare disease tiering 2025 [updated 19 September 2025. Available from: https://re-docs.genomicsengland.co.uk/tiering/.

20. Travasci S. Simple-icd-10 2.1.1. 2025.

21. Schlicker A, Domingues FS, Rahnenführer J, Lengauer T. A new measure for functional similarity of gene products based on Gene Ontology. BMC Bioinformatics. 2006;7:302.

22. Murtagh F, Legendre P. Ward’s hierarchical clustering method: clustering criterion and agglomerative algorithm. arXiv preprint arXiv:11116285. 2011.

23. England N. Primary Reason for Referral (Mental Health). NHS Data Model and Dictionary2024.

24. Stiebahl S. Obesity statistics. In: Library HoC, editor. 2025.

25. Egli V, Hobbs M, Carlson J, Donnellan N, Mackay L, Exeter D, et al. Deprivation matters: understanding associations between neighbourhood deprivation, unhealthy food outlets, unhealthy dietary behaviours and child body size using structural equation modelling. Journal of Epidemiology and Community Health. 2020;74(5):460–6.

26. Kim Y, Cubbin C, Oh S. A systematic review of neighbourhood economic context on child obesity and obesity-related behaviours. Obesity Reviews. 2019;20(3):420–31.

27. Staatz CB, Kelly Y, Lacey RE, Hardy R. Area-level and family-level socioeconomic position and body composition trajectories: longitudinal analysis of the UK Millennium Cohort Study. The Lancet Public Health. 2021;6(8):e598–e607.

28. White J, Rehkopf D, Mortensen HL. Trends in Socioeconomic Inequalities in Body Mass Index, Underweight and Obesity among English Children, 2007–2008 to 2011–2012. PLOS ONE. 2016;11(1):e0147614.

29. Sudlow C, Gallacher J, Allen N, Beral V, Burton P, Danesh J, et al. UK Biobank: An Open Access Resource for Identifying the Causes of a Wide Range of Complex Diseases of Middle and Old Age. PLOS Medicine. 2015;12(3):e1001779.

30. Corrochano S, Männikkö R, Joyce IP, Mcgoldrick P, Wettstein J, Lassi G, et al. Novel mutations in human and mouse SCN4A implicate AMPK in myotonia and periodic paralysis. Brain. 2014;137(12):3171–85.

31. Kolk DVWB, Saari S, Lovric A, Arif M, Alvarez M, Ko A, et al. Molecular pathways behind acquired obesity: Adipose tissue and skeletal muscle multiomics in monozygotic twin pairs discordant for BMI. Cell Reports Medicine. 2021;2(4):100226.

32. Stoetzel C, Laurier V, Davis EE, Muller J, Rix S, Badano LJ, et al. BBS10 encodes a vertebrate-specific chaperonin-like protein and is a major BBS locus. Nature Genetics. 2006;38(5):521–4.

33. Zaghloul AN, Katsanis N. Mechanistic insights into Bardet-Biedl syndrome, a model ciliopathy. Journal of Clinical Investigation. 2009;119(3):428–37.

34. Tanaka JA, Cho TM, Retterer K, Jones RJ, Nowak C, Douglas J, et al. De novo pathogenic variants in *CHAMP1* are associated with global developmental delay, intellectual disability, and dysmorphic facial features. Molecular Case Studies. 2016;2(1):a000661.

35. Garrity M, Kavus H, Rojas-Vasquez M, Valenzuela I, Larson A, Reed S, et al. Neurodevelopmental phenotypes in individuals with pathogenic variants in *CHAMP1*. Molecular Case Studies. 2021;7(4):a006092.

36. Stekelenburg C, Blouin J-L, Santoni F, Zaghloul N, O’Hare AE, Dusaulcy R, et al. Loss of Nexmif results in the expression of phenotypic variability and loss of genomic integrity. Scientific Reports. 2022;12(1).

37. Langley E, Farach SL, Koenig KM, Northrup H, Rodriguez-Buritica FD, Mowrey K. *NEXMIF* pathogenic variants in individuals of Korean, Vietnamese, and Mexican descent. American Journal of Medical Genetics Part A. 2022;188(6):1688–92.

38. Anastasescu MC, Gheorman V, Godeanu VS, Cojocaru A, Iliuta PF, Stepan DM, et al. KIAA2022/NEXMIF c.1882C>T (p.Arg628*) Variant in a Romanian Patient with Neurodevelopmental Disorders and Epilepsy: A Case Report and Systematic Review. Life. 2025;15(3):497.

39. Yadav H, Quijano C, Kamaraju KA, Gavrilova O, Malek R, Chen W, et al. Protection from Obesity and Diabetes by Blockade of TGF-β/Smad3 Signaling. Cell Metabolism. 2011;14(1):67–79.

40. Huvenne H, Dubern B, Clément K, Poitou C. Rare Genetic Forms of Obesity: Clinical Approach and Current Treatments in 2016. Obesity Facts. 2016;9(3):158–73.

41. Kessler J, Koebnick C, Smith N, Adams A. Childhood Obesity Is Associated With Increased Risk of Most Lower Extremity Fractures. Clinical Orthopaedics & Related Research. 2013;471(4):1199–207.

42. Patel R, Gillis G, Mackay CE, Griffanti L, Wang C, Ebmeier KP, et al. The lifetime accumulation of multimorbidity and its influence on dementia risk: a UK Biobank study. Brain Communications. 2025;7(4).

43. Smail A, Al-Jawahiri R, Baker K. Polycomb-associated and Trithorax-associated developmental conditions—phenotypic convergence and heterogeneity. European Journal of Human Genetics. 2025;33(11):1414–21.

