## Supplementary Material for "Young people with obesity and rare disease – genotypes, phenotypes and healthcare use"

[Table S3: Genes enriched in case set (p_raw_ <0.01)	4](#_Toc237708455)

### Table S1: HPO and ICD-10 codes applied for case definition

| **HPO Terms (Version: 2.0.6)** | **ICD-10 Codes (Version: 2019)*** |
| --- | --- |
| 1. HP:0001513 - Obesity 2. HP:0025501 - Class III obesity 3. HP:0025500 - Class II obesity 4. HP:0025499 - Class I obesity 5. HP:0012743 - Abdominal obesity 6. HP:0008915 - Childhood-onset truncal obesity 7. HP:0001956 - Truncal obesity | 1. E66 - Obesity 2. E66.0 - Obesity due to excess calories 3. E66.2 - Extreme obesity with alveolar hypoventilation 4. E66.8 Other obesity 5. E66.9 Obesity, unspecified |

*ICD-10 code “E66.1 Drug-induced obesity” was not included in our filtering criteria

### Table S2: Obesity-related terms in HPO phenotype source cases

| Obesity-related HPO terms | No. of Patients |
| --- | --- |
| One | 229 |
| Multiple | 35 |
| Distribution of Terms |  |
| HP:001513 - Obesity | 205 |
| HP:001956 - Truncal obesity or HP:0012743 - Abdominal/central obesity | 77 |
| HP:0008915 - Childhood-onset truncal obesity | 17 |

### Table S3: Genes enriched in case set (p_raw_ <0.01)

| Gene | Case Count | Control Count | Odds Ratio | Raw p-value* | Adj p-value (BH) |
| --- | --- | --- | --- | --- | --- |
| *SCN4A* | 3 | 1 | 69.1 | 2.83E-04 | 0.113 |
| *FLNB* | 7 | 27 | 6.00 | 4.34E-04 | 0.113 |
| *SIL1* | 2 | 0 | *Inf* | 1.75E-03 | 0.182 |
| *MC4R* | 2 | 0 | *Inf* | 1.75E-03 | 0.182 |
| *BBS10* | 2 | 0 | *Inf* | 1.75E-03 | 0.182 |
| *BMPR1B* | 3 | 5 | 13.8 | 3.49E-03 | 0.182 |
| *IARS2* | 2 | 1 | 46.0 | 5.10E-03 | 0.315 |
| *PIK3CD* | 2 | 1 | 46.0 | 5.10E-03 | 0.315 |
| *PITRM1* | 13 | 127 | 2.38 | 5.85E-03 | 0.315 |
| *NEXMIF* | 8 | 58 | 3.19 | 6.06E-03 | 0.315 |
| *CHAMP1* | 4 | 15 | 6.14 | 7.12E-03 | 0.334 |
| *COL4A4* | 3 | 8 | 8.63 | 9.36E-03 | 0.334 |
| *QARS* | 2 | 2 | 23.0 | 9.92E-03 | 0.334 |
| *TGFB2* | 2 | 2 | 23.0 | 9.92E-03 | 0.334 |
| *GMPPA* | 2 | 2 | 23.0 | 9.92E-03 | 0.334 |

* One-tailed test

### Table S4: Top-level HPO terms enriched or depleted in cases versus controls

| HPO Term | % of case | % of control | Delta % | p_raw_ | p_adj_ (BH) |
| --- | --- | --- | --- | --- | --- |
| Abnormality of the nervous system | 85.4 | 69.23 | 16.17 | 1.21E-14 | 2.78E-13^***^ |
| Abnormality of the endocrine system | 11.6 | 4.65 | 6.95 | 2.20E-12 | 2.53E-11^***^ |
| Growth abnormality | 33 | 21.57 | 11.43 | 1.57E-09 | 1.20E-08^***^ |
| Abnormality of the breast | 3.8 | 1.07 | 2.73 | 3.73E-08 | 2.14E-07^***^ |
| Abnormality of the genitourinary system | 21.2 | 15.7 | 5.5 | 1.01E-03 | 4.65E-03^**^ |
| Abnormality of limbs | 27.2 | 21.16 | 6.04 | 1.26E-03 | 4.82E-03^**^ |
| Abnormality of the cardiovascular system | 13.2 | 18.12 | -4.92 | 4.96E-03 | 1.63E-02^*^ |
| Abnormality of the integument | 23.8 | 19.49 | 4.31 | 1.78E-02 | 5.11E-02 |
| Abnormality of head or neck | 45.6 | 41.52 | 4.08 | 7.04E-02 | 1.80E-01 |
| Abnormality of the digestive system | 16.4 | 19.43 | -3.03 | 9.25E-02 | 2.13E-01 |
| Abnormality of metabolism/homeostasis | 12 | 10 | 2 | 1.45E-01 | 3.04E-01 |
| Abnormality of the voice | masked*^#^* | masked*^#^* | -0.58 | 1.87E-01 | 3.58E-01 |
| Neoplasm | 1.8 | 2.44 | -0.64 | 3.63E-01 | 6.42E-01 |
| Abnormality of the eye | 26.4 | 28 | -1.6 | 4.36E-01 | 7.16E-01 |
| Abnormality of prenatal development or birth | 7.8 | 8.55 | -0.75 | 5.54E-01 | 8.50E-01 |
| Abnormality of the respiratory system | 8.6 | 9.3 | -0.7 | 5.99E-01 | 8.60E-01 |
| Abnormality of blood and blood-forming tissues | 3.8 | 4.23 | -0.43 | 6.40E-01 | 8.66E-01 |
| Abnormality of the musculoskeletal system | 50.8 | 51.65 | -0.85 | 7.09E-01 | 8.74E-01 |
| Abnormality of the ear | 16.8 | 16.2 | 0.6 | 7.22E-01 | 8.74E-01 |
| Abnormality of the immune system | 10.8 | 11.17 | -0.37 | 7.98E-01 | 9.18E-01 |
| Constitutional symptom | 2.8 | 2.85 | -0.05 | 9.49E-01 | 1.00E+00 |
| Abnormal cellular phenotype | 1.2 | 1.2 | 0 | 9.95E-01 | 1.00E+00 |
| Abnormality of the thoracic cavity | masked*^#^* | masked*^#^* | -0.02 | 1.00E+00 | 1.00E+00 |

*\# Terms associated with fewer than 5 patients in either cohort have been masked to protect patient privacy*

**** p_adj_ <0.001, ** p_adj_ <0.01, * p_adj_ <0.05*

### Table S5: Propagated HPO terms enriched or depleted in cases versus controls

| HPO Term^##^ | % of case | % of control | Delta % | p_raw_ | p_adj_ (BH) |
| --- | --- | --- | --- | --- | --- |
| Polyphagia | 5.8 | 0.05 | 5.75 | 8.23E-120 | 4.82E-116^***^ |
| Abnormal consumption behaviour | 9.6 | 0.52 | 9.08 | 8.94E-98 | 2.62E-94^***^ |
| Abnormal eating behaviour | 9 | 0.45 | 8.55 | 1.87E-96 | 3.65E-93^***^ |
| Tall stature | 16.2 | 2.47 | 13.73 | 2.06E-68 | 3.02E-65^***^ |
| Tapered finger | 3.8 | 0.38 | 3.42 | 1.81E-25 | 2.12E-22^***^ |
| Recurrent maladaptive behaviour | 16 | 5.14 | 10.86 | 4.34E-25 | 4.24E-22^***^ |
| Increased body weight | 6 | 1.04 | 4.96 | 1.35E-22 | 1.13E-19^***^ |
| Abnormality of mental function | 73.6 | 52.09 | 21.51 | 3.96E-21 | 2.90E-18^***^ |
| Atypical behaviour | 35.2 | 19.44 | 15.76 | 7.57E-18 | 4.92E-15^***^ |
| Abnormal nervous system physiology | 83.4 | 65.67 | 17.73 | 2.08E-16 | 1.22E-13^***^ |
| Sleep abnormality | 9.4 | 2.98 | 6.42 | 2.25E-15 | 1.10E-12^***^ |
| Abnormality of body height | 25.8 | 13.39 | 12.41 | 4.09E-15 | 1.84E-12^***^ |
| Abnormality of the nervous system | 85.4 | 69.23 | 16.17 | 1.21E-14 | 5.05E-12^***^ |
| Sleep apnoea | 3.8 | 0.72 | 3.08 | 1.70E-13 | 6.22E-11^***^ |
| Sleep-related breathing disorders | 3.8 | 0.74 | 3.06 | 5.91E-13 | 1.82E-10^***^ |
| Abnormality of the endocrine system | 11.6 | 4.65 | 6.95 | 2.20E-12 | 6.13E-10^***^ |
| Intellectual disability | 55.6 | 40.69 | 14.91 | 3.45E-11 | 8.42E-09^***^ |
| Long fingers | 4.4 | 1.16 | 3.24 | 3.81E-10 | 6.75E-08^***^ |
| Neurodevelopmental abnormality | 71.6 | 57.62 | 13.98 | 5.50E-10 | 9.48E-08^***^ |
| Growth abnormality | 33 | 21.57 | 11.43 | 1.57E-09 | 2.48E-07^***^ |
| Abnormality of the genital system | 10 | 5.02 | 4.98 | 9.45E-07 | 9.54E-05^***^ |
| Neurodevelopmental delay | 61.4 | 50.59 | 10.81 | 2.23E-06 | 2.14E-04^***^ |
| Abnormal external genitalia morphology | 9 | 4.58 | 4.42 | 5.54E-06 | 5.07E-04^***^ |
| Abnormal male external genitalia morphology | 8.6 | 4.33 | 4.27 | 6.86E-06 | 6.18E-04^***^ |
| Abnormality of the male genitalia | 8.6 | 4.43 | 4.17 | 1.33E-05 | 1.17E-03^**^ |
| Hypertonia | Masked^#^ | Masked^#^ | -3.44 | 1.38E-05 | 1.19E-03^**^ |
| Abnormal reproductive system morphology | 9.2 | 4.88 | 4.32 | 1.65E-05 | 1.40E-03^**^ |
| Abnormality of limb bone | 19.8 | 13.14 | 6.66 | 1.91E-05 | 1.58E-03^**^ |
| Abnormal limb bone morphology | 19.8 | 13.14 | 6.66 | 1.91E-05 | 1.58E-03^**^ |
| Abnormality of the hand | 15.8 | 10.12 | 5.68 | 4.48E-05 | 3.45E-03^**^ |
| Abnormal appendicular skeleton morphology | 22 | 15.33 | 6.67 | 5.71E-05 | 4.28E-03^**^ |
| Macrocephaly | 9 | 4.96 | 4.04 | 6.22E-05 | 4.56E-03^**^ |
| Increased head circumference | 9 | 4.96 | 4.04 | 6.22E-05 | 4.56E-03^**^ |
| Abnormality of the upper limb | 17.4 | 11.58 | 5.82 | 7.92E-05 | 5.66E-03^**^ |
| Abnormality of the musculature | 14.4 | 21.69 | -7.29 | 1.00E-04 | 6.89E-03^**^ |
| Autistic behaviour | 21.2 | 14.9 | 6.3 | 1.20E-04 | 8.00E-03^**^ |
| Diagnostic behavioural phenotype | 21.2 | 14.9 | 6.3 | 1.20E-04 | 8.00E-03^**^ |
| Abnormal muscle tone | 10.2 | 16.22 | -6.02 | 3.23E-04 | 1.99E-02^*^ |
| Abnormal cognitive process | 42 | 34.58 | 7.42 | 6.52E-04 | 3.54E-02^*^ |
| Abnormal communication | 42 | 34.58 | 7.42 | 6.52E-04 | 3.54E-02^*^ |
| Abnormal language feature | 41.8 | 34.45 | 7.35 | 7.35E-04 | 3.88E-02^*^ |
| Abnormal speech pattern | 41.8 | 34.45 | 7.35 | 7.35E-04 | 3.88E-02^*^ |
| Abnormal muscle physiology | 12.6 | 18.54 | -5.94 | 7.68E-04 | 3.98E-02^*^ |
| Global developmental delay | 47.2 | 39.82 | 7.38 | 9.83E-04 | 4.88E-02^*^ |
| Language impairment | 41 | 33.86 | 7.14 | 9.84E-04 | 4.88E-02^*^ |
| Motor delay | 36 | 29.13 | 6.87 | 9.73E-04 | 4.88E-02^*^ |
| Abnormality of the genitourinary system | 21.2 | 15.7 | 5.5 | 1.01E-03 | 4.97E-02^*^ |

*# Terms associated with fewer than 5 patients in either group have been masked to protect patient privacy*

*## Terms listed if* |Δ|>3% and p_adj_ <0.05

**** p_adj_ <0.001, ** p_adj_ <0.01, * p_adj_ <0.05*

### Table S6: Top 10 primary diagnoses within case and control groups - all admissions per patient

| ICD-10 | Term | Test | Odds Ratio | Raw p-value | Adj p-value (BH) |
| --- | --- | --- | --- | --- | --- |
| N185 | Chronic kidney disease, stage 5 | Chi² | 5.79 | <0.001 | **<0.001***** |
| D619 | Aplastic anaemia, unspecified | Chi² | 5.90 | 6.55E-61 | **4.92E-60***** |
| J22X | Unspecified acute lower respiratory infection | Chi² | 0.59 | 1.65E-05 | **3.54E-05***** |
| G473 | Sleep apnoea | Chi² | 1.55 | 1.62E-03 | **2.43E-03***** |
| E301 | Precocious puberty | Chi² | 6.11 | 8.66E-36 | **4.33E-35***** |
| E230 | Hypopituitarism | Chi² | 3.11 | 4.27E-12 | **1.28E-11***** |
| G403 | Generalised idiopathic epilepsy and epileptic syndromes | Chi² | 0.87 | 0.390 | 0.450 |
| K590 | Constipation | Chi² | 0.81 | 0.203 | 0.277 |
| Q780 | Osteogenesis Imperfecta | Chi² | 0.95 | 0.758 | 0.813 |
| K029 | Dental caries, unspecified | Chi² | 0.97 | 0.858 | 0.858 |
| N390 | Urinary tract infection, site not specified | Chi² | 0.84 | 0.312 | 0.390 |
| J069 | Acute upper respiratory infection, unspecified | Chi² | 0.52 | 4.16E-04 | **7.79E-04***** |
| B349 | Viral infection, unspecified | Chi² | 0.53 | 6.94E-04 | **1.16E-03***** |
| G409 | Epilepsy, unspecified | Chi² | 0.36 | 1.07E-06 | **2.68E-06***** |
| Z755 | Holiday relief care | Fisher | 0 | 1.73E-20 | **6.48E-20***** |

*^#^ Between 31/07/2017 - 31/07/2022*

**** p_adj_ <0.001, ** p_adj_ <0.01, * p_adj_ <0.05.*

### Table S7: Top 10 inpatient diagnoses within case and control groups - unique per patient

| ICD-10 | Term | Test | Odds Ratio | Raw p-value | Adj p-value (BH) |
| --- | --- | --- | --- | --- | --- |
| G473 | Sleep apnoea | Chi² | 2.16 | 3.27E-05 | **1.42E-04***** |
| K029 | Dental caries, unspecified | Chi² | 1.29 | 0.176 | 0.287 |
| E669 | Acute upper respiratory infection, unspecified | Chi² | 0.78 | 0.263 | 0.380 |
| J069 | Obesity, unspecified | Fisher | - | *NA* | *NA* |
| J22X | Unspecified acute lower respiratory infection | Chi² | 0.55 | 6.91E-03 | **0.0225*** |
| B349 | Viral infection, unspecified | Chi² | 0.68 | 0.0911 | 0.197 |
| K590 | Constipation | Chi² | 1.21 | 0.425 | 0.503 |
| R568 | Other and unspecified convulsions | Chi² | 0.82 | 0.416 | 0.503 |
| A099 | Gastroenteritis and colitis of unspecified origin | Chi² | 0.97 | 0.902 | 0.902 |
| E230 | Hypopituitarism | Chi² | 3.60 | 8.15E-07 | **5.29E-06***** |
| J039 | Acute tonsillitis, unspecified | Chi² | 0.84 | 0.531 | 0.576 |
| G409 | Epilepsy, unspecified | Chi² | 0.55 | 0.0360 | 0.0937 |
| R11X | Vomiting, unspecified | Chi² | 0.66 | 0.143 | 0.266 |

*^#^ Between 31/07/2017 - 31/07/2022*

**** p_adj_ <0.001, ** p_adj_ <0.01, * p_adj_ <0.05.*

### Table S8: HPO terms enriched within clusters

| HPO Term ^#^ | Enrichment Cluster | OR | P_raw_ | p_adj_ (BH) |
| --- | --- | --- | --- | --- |
| Global developmental delay | 1 | 8.17 | 6.20E-20 | 1.42E-17^***^ |
| Intellectual disability | 1 | 4.16 | 1.91E-10 | 3.49E-08^***^ |
| Delayed fine motor development | 1 | 23.1 | 5.46E-31 | 2.50E-28^***^ |
| Autistic behaviour | 1 | 5.23 | 1.02E-07 | 1.55E-05^***^ |
| Intellectual disability, moderate | 1 | 5.57 | 1.32E-04 | 1.21E-02^*^ |
| Delayed speech and language development | 1 | 14.2 | 1.03E-28 | 3.14E-26^***^ |
| Inability to walk | 1 | *Inf* | 5.70E-05 | 5.80E-03^**^ |
| Delayed gross motor development | 1 | 24.3 | 1.21E-33 | 1.11E-30^***^ |
| Tall stature | 2 | 16.3 | 2.89E-12 | 1.32E-09^***^ |
| Truncal obesity | 2 | 6.16 | 5.72E-06 | 1.05E-03^**^ |
| Autism | 2 | 6.35 | 2.52E-04 | 3.29E-02^*^ |
| Polyphagia | 2 | 34.3 | 5.95E-13 | 5.44E-10^***^ |
| Recurrent long bone fractures | 6 | 171 | 4.64E-05 | 2.12E-02^*^ |
| Prominent forehead | 6 | 85 | 1.15E-04 | 3.49E-02^*^ |
| Osteopenia | 6 | *Inf* | 1.18E-05 | 1.08E-02^*^ |
| Visual impairment | 7 | 52.2 | 3.48E-06 | 1.59E-03^**^ |
| Nystagmus | 7 | 44.8 | 3.97E-05 | 1.21E-02^*^ |
| Septo-optic dysplasia | 7 | 426 | 1.74E-08 | 1.59E-05^***^ |
| Abnormality of the respiratory system | 9 | *Inf* | 7.84E-06 | 7.17E-03^**^ |
| Hydronephrosis | 10 | *Inf* | 4.90E-06 | 4.48E-03^**^ |

*# Terms listed if* OR>1 and p_adj_ <0.05 compared to remainder

**** p_adj_ <0.001, ** p_adj_ <0.01, * p_adj_ <0.05*

### Table S9: Overlap in gene-sets observed within phenotypic clusters

| Cluster Pair* | | No. of shared genes | Jaccard index | Shared genes |
| --- | --- | --- | --- | --- |
| 1 | 2 | 29 | 0.099 | *AFF4, ATP7A, BCOR, CCDC22, CHD7, CLTC, CREBBP, EHMT1, FAT4, FLNA, HIVEP2, IARS2, KAT6A, KCNQ2, KIDINS220, KMT2D, MACF1, MBTPS2, MECP2, NHS, PDE4D, PDHA1, PITRM1, RANBP2, SETD2, SHANK3, SKI, SLC9A6, ZSWIM6* |
| 1 | 6 | 3 | 0.011 | *FLNA, STAG1, TRIO* |
| 1 | 7 | 15 | 0.053 | *BCOR, CHD3, DEAF1, DMD, EHMT1, EP300, FMR1, KMT2C, KMT2D, NLGN3, PTCH1, SPG11, STAG1, THOC2, TSC2* |
| 1 | 9 | 4 | 0.015 | *CDKL5, KIF1A, PKD1, TSC2* |
| 1 | 10 | 3 | 0.011 | *DCHS1, MACF1, SPECC1L* |
| 2 | 6 | 1 | 0.017 | *FLNA* |
| 2 | 7 | 3 | 0.038 | *BCOR, EHMT1, KMT2D* |
| 2 | 10 | 1 | 0.016 | *MACF1* |
| 6 | 7 | 1 | 0.029 | *STAG1* |
| 7 | 9 | 1 | 0.026 | *TSC2* |

**Cluster pairs not tabulated had zero shared genes*

### Figure S1: 100KGP disease recruitment categories for cases and controls

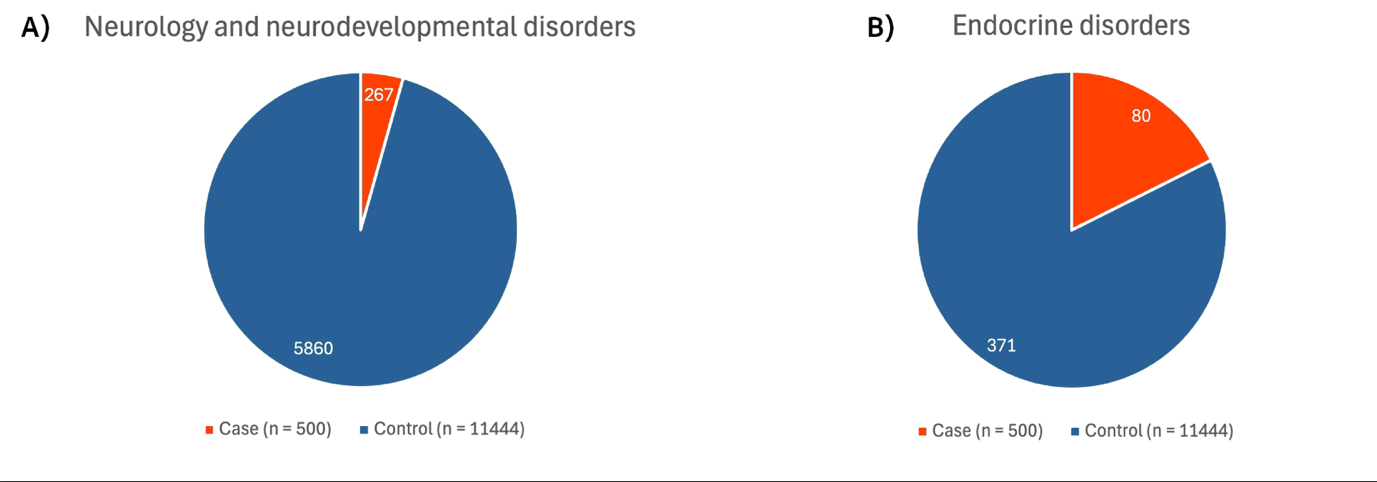

A) Proportion of cases and controls within neurology and neurodevelopmental recruitment category.

B) Proportion of cases and controls within endocrine recruitment category.

### Figure S2: Demographic characteristics of case and control groups

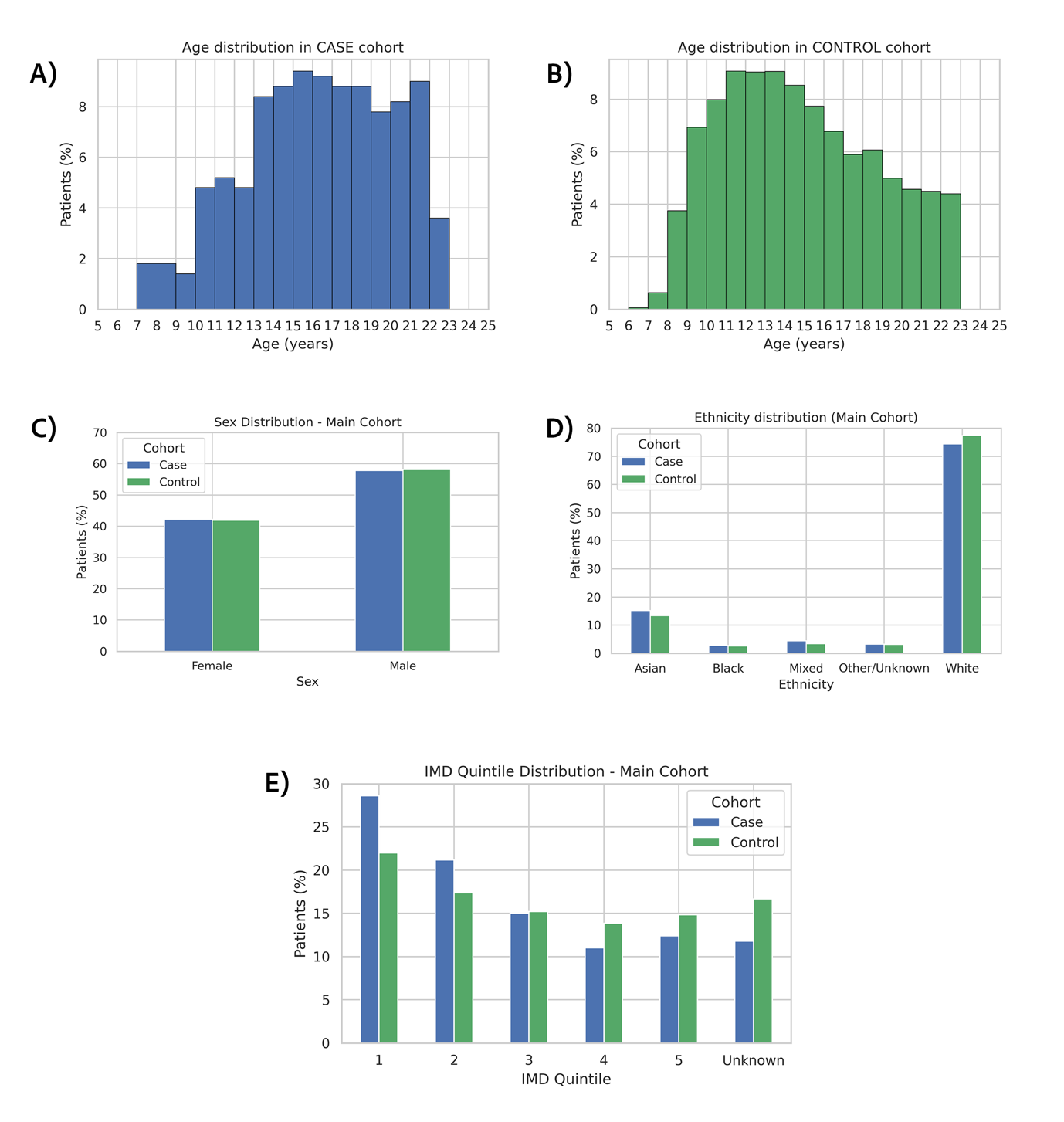

A) Age distribution in case cohort. Bins containing less than 5 patients have been merged with a neighbouring bin to protect patient privacy.

B) Age distribution in control cohort.

C) Sex distribution in both cohorts.

D) Ethnicity distribution in both cohorts.

E) IMD quintile distribution in both cohorts.

### Figure S3: HPO term counts in case and control groups

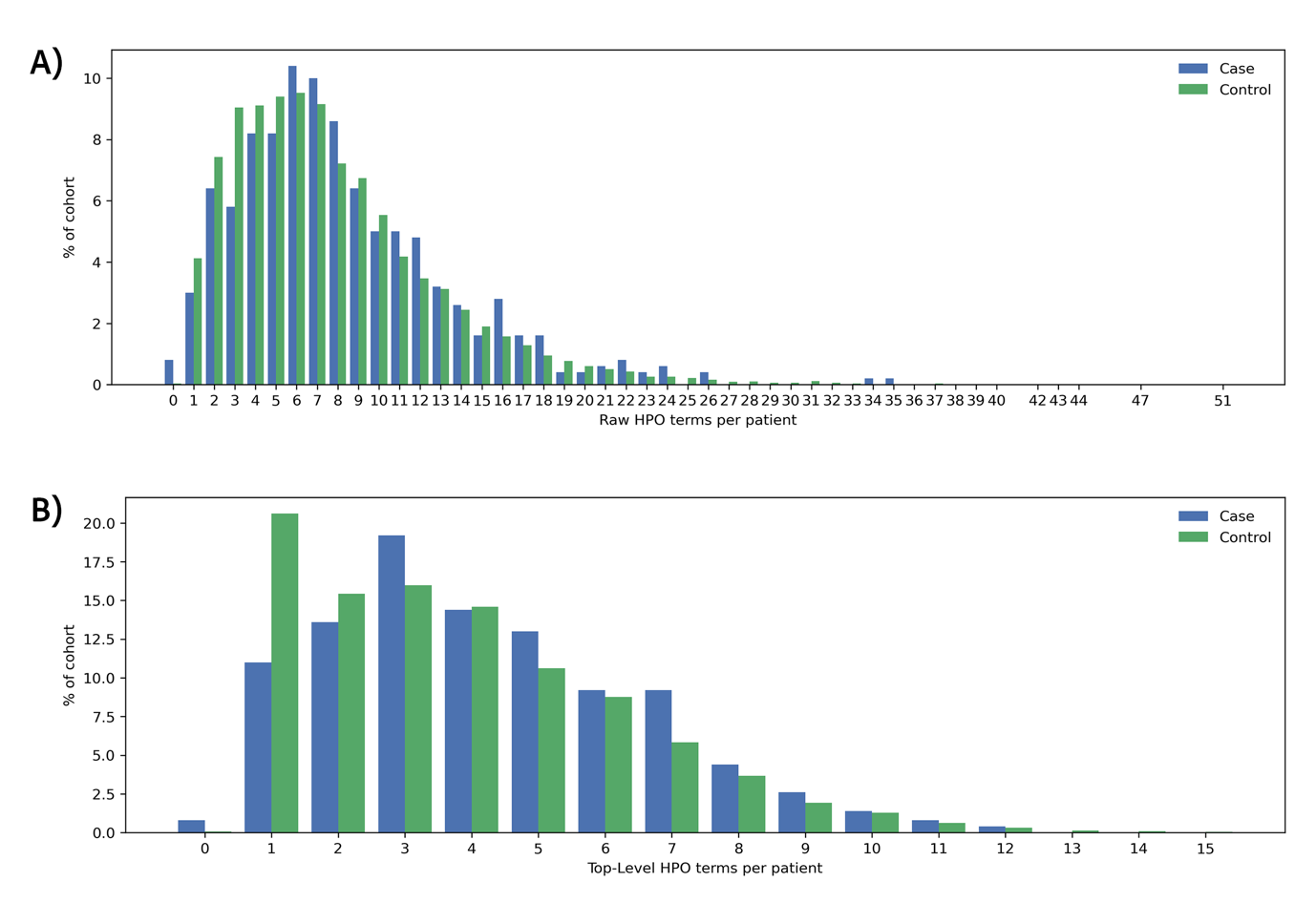

A) Distribution of raw clinician-reported HPO terms per patient.

B) Distribution of propagated top-level (organ system-level) HPO terms per patient.

### Figure S4: A&E visit counts, chief complaints and primary diagnoses

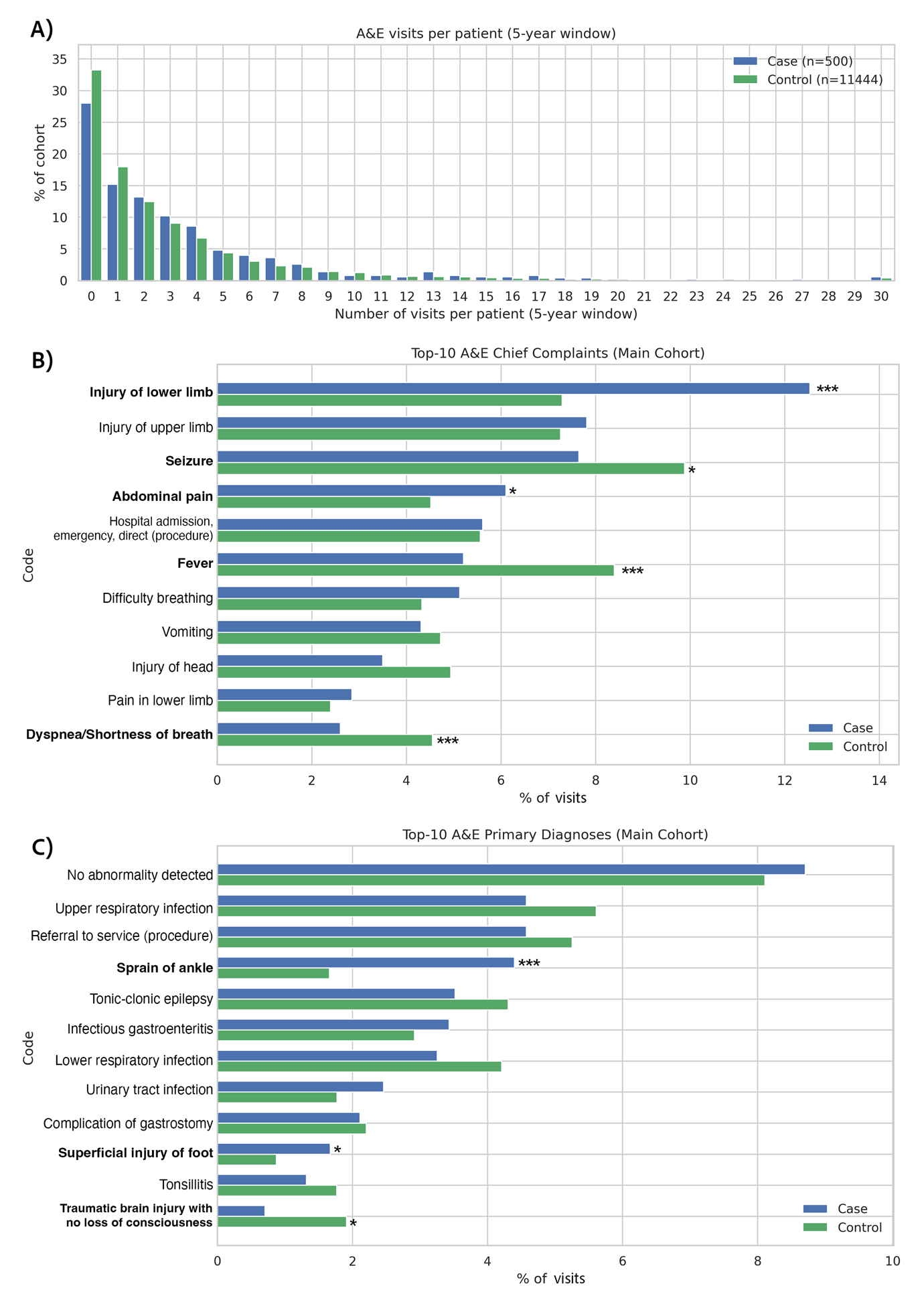

A) Distribution of A&E visits per patient (31/07/2017 - 31/07/2022). Patients with 30 or more visits are grouped in the “30” bin.

B) Top 10 chief complaints.

C) Top 10 primary diagnoses

**** p_adj_ <0.001, ** p_adj_ <0.01*, * p_adj_ <0.05

### Figure S5: Inpatient admission counts and primary diagnoses

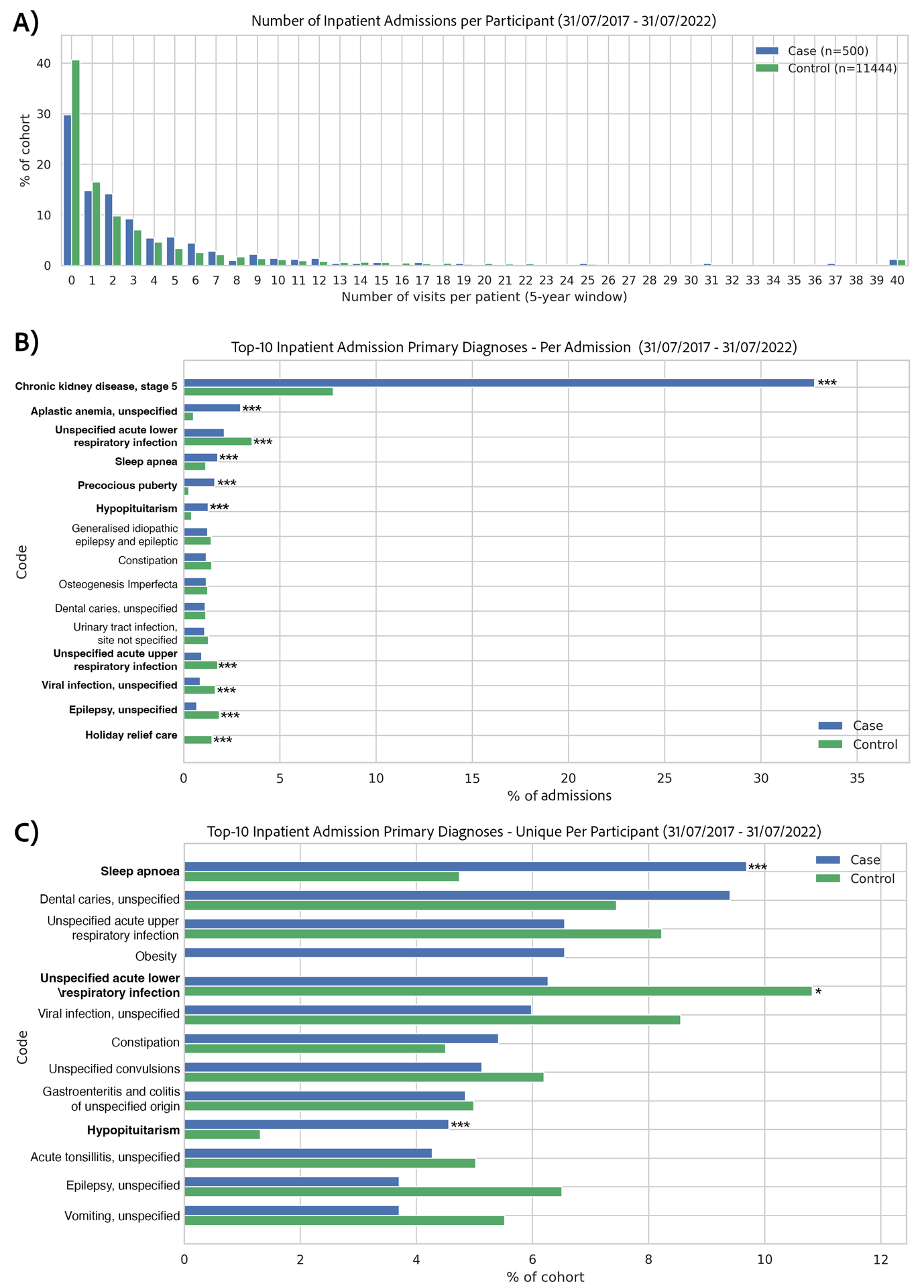

A) Distribution of admissions per patient (31/07/2017 - 31/07/2022). Patients with 40 or more admissions are grouped in the “40” bin.
B) Top 10 Primary Admission Reasons – Per Admission.
C) Top 10 Primary Admission Reasons – Per Patient.

**** p_adj_ <0.001, ** p_adj_ <0.01, * p_adj_ <0.05.*

### Figure S6: Obesity phenotype clustering diagnostic plots

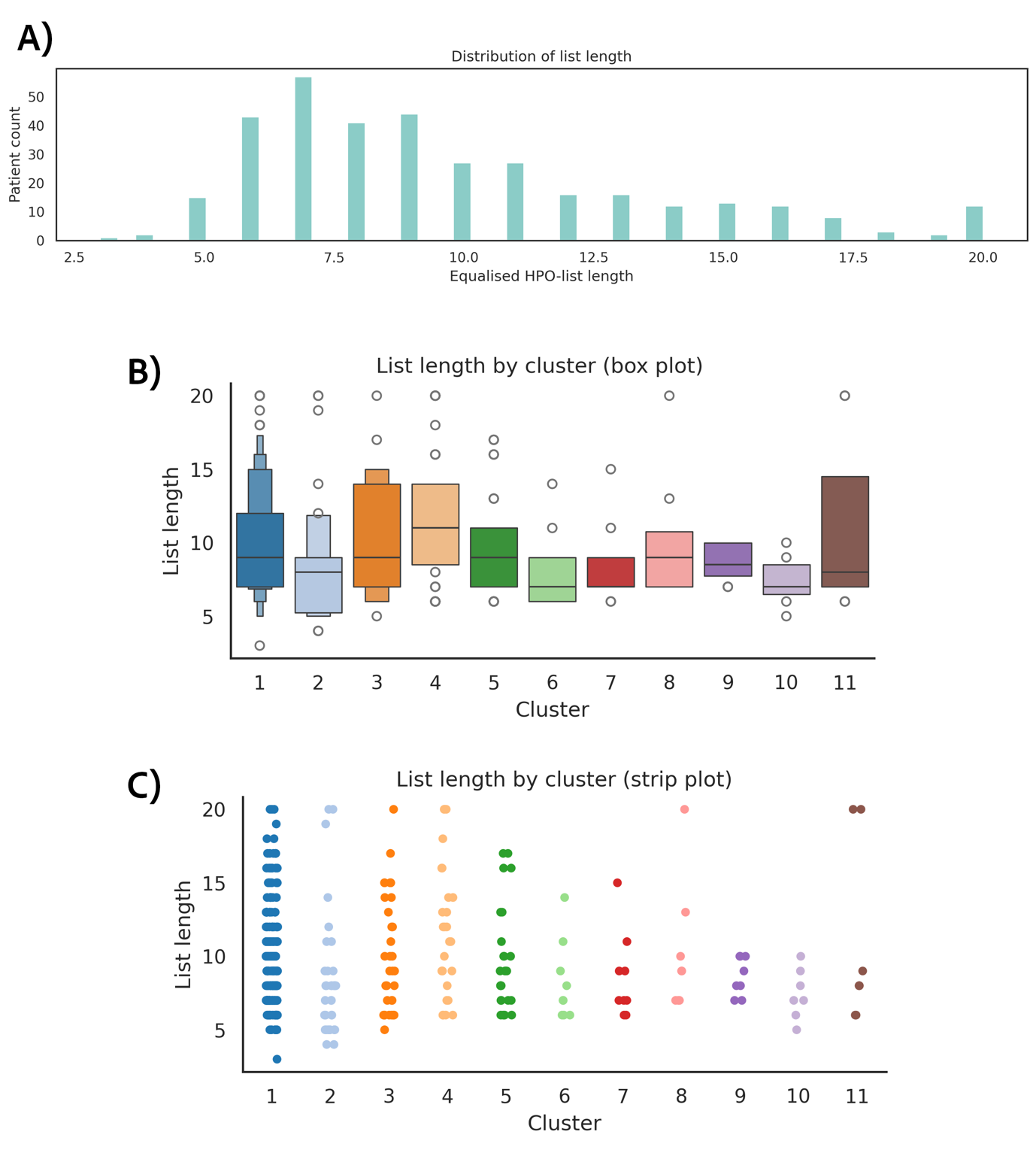

A) Bar chart showing the number of patients with each HPO list length after equalisation.

B) Box plot showing the median and quartile of HPO list length by patient cluster.

C) Scatter plot showing the distribution of HPO list length by patient cluster. All plots show high intra and inter-cluster variability and indicate that list length is not the key factor driving cluster formation.

### Figure S7: Within-cluster gene ontology networks

Cluster 1 (n=184)

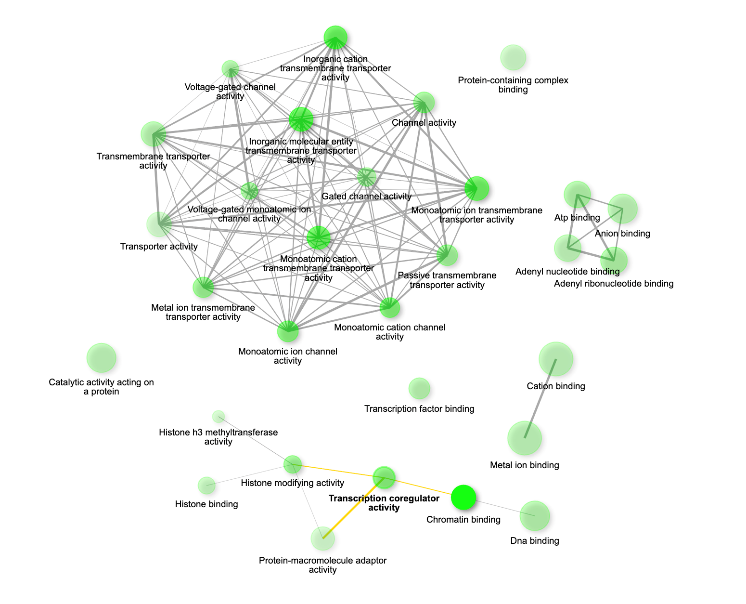

Cluster 2 (n=34)

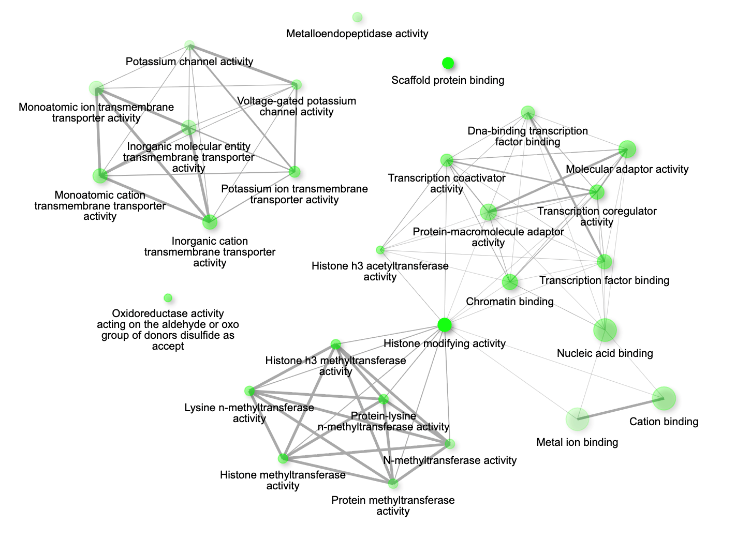

Cluster 7 (n=9)

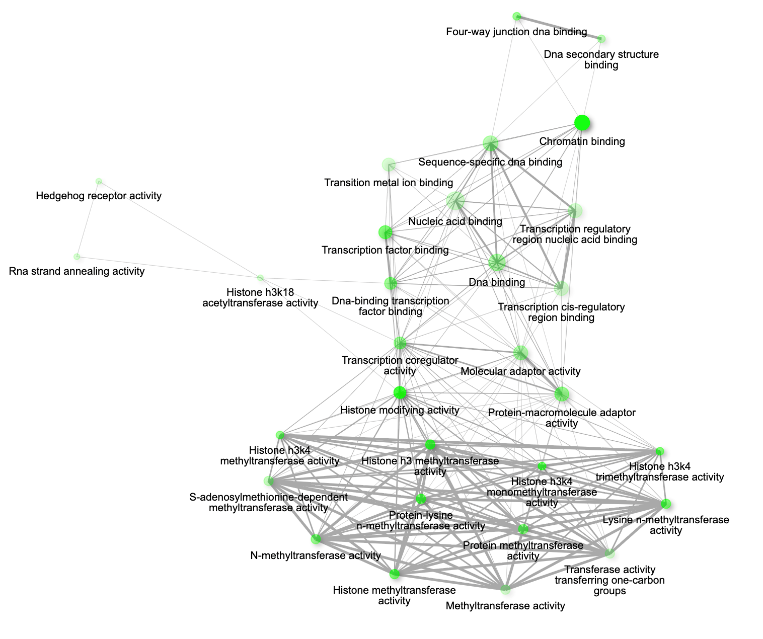
